# A nonrandomized phase 2 trial of oral thymic peptides in hospitalized patients with Covid-19

**DOI:** 10.1101/2021.12.05.21267318

**Authors:** Héctor M. Ramos-Zaldívar, Karla G. Reyes-Perdomo, Nelson A. Espinoza-Moreno, Ernesto Tomás Dox-Cruz, Thania Camila Aguirre Urbina, Astrid Yohaly Rivera Caballero, Eduardo Smelin Perdomo Dominguez, Sofía Guadalupe Peña Calix, Joselin Michelle Monterroso-Reyes, Erick Fernando Caballero Vásquez, Tarek Sai Zelaya Ortiz, Hilbron Eduardo Rodríguez-Machado, Marcelo Andres Forgas Solis, Iveth Sebilla Silva, Mauricio Edgardo Zavala Galeano, Alejandro Antonio Morga Alvarado, Angie María Nicolle Solís Medina, Leticia M. Guerrero-Díaz, Julia E. Jiménez-Faraj, Caroll Alejandra Perelló Santos, Wilberg A. Moncada Arita, Darwing Fabricio Valdiviezo Montufar, Josué David Hernández Sabillón, Mónica L. Sorto G., Xochilt Xiomara Padilla Navarro, Victoria A. Palomo-Bermúdez, Héctor Armando Alvarenga Andino, Sandra Patricia Reyes Guzman, María Haydee Rivera Reyes, Esdras Said Medina Paz, Joselyn Rosario Alvarado Enamorado, Yenny Mariel Sabillón Sagastume, Ariadna Stephanny Mejia Rivera, Claudia Michelle Posas Sarmiento, Xenia Vanessa Jiménez Pineda, Verónica Alejandra Hernández Puerto, Josué David Portillo Landaverde, Reyes S. Sergio, Ivin Perdomo R., Josué J. Rivera, Wendy Cecilia Mendoza Girón, Karla Melissa Tróchez Sabillón, Paola Nohemy Katsumata Leiva, Karla Elizabeth Pineda Toro, Jimena A. Montes-Gambarelli, Cristhiam Flores, Edison Salas-Huenuleo, Marcelo E. Andia

**Author notes:** Corresponding author: Héctor Miguel Ramos Zaldívar, M.D., Contact information: Primary, Secondary.

## Abstract

**Background:** Coronavirus disease 2019 (Covid-19) active cases continue to demand the development of safe and effective treatments. This is the first clinical trial to evaluate the safety and efficacy of oral thymic peptides.

**Methods:** We conducted a nonrandomized phase 2 trial with a historic control group to evaluate the safety and efficacy of a daily 250-mg oral dose of thymic peptides in the treatment of hospitalized Covid-19 patients. Comparison based on standard care from registry data was performed after propensity score matching. The primary outcomes were survival, time to recovery and the number of participants with treatment-related adverse events or side effects by day 20.

**Results:** A total of 44 patients were analyzed in this study, 22 in the thymic peptides group and 22 in the standard care group. There were no deaths in the intervention group, compared to 24% mortality in standard care by day 20 (log-rank P=0.02). The Kaplan-Meier analysis showed a significantly shorter time to recovery by day 20 in the thymic peptides group as compared with standard care (median, 6 days vs. 12 days; hazard ratio for recovery, 2.75 [95% confidence interval, 1.34 to 5.62]; log-rank P=0.002). No side effects or adverse events were reported.

**Conclusion:** In patients hospitalized with Covid-19, the use of thymic peptides reported no side effects, adverse events, or deaths by day 20. When compared with registry data, a significantly shorter time to recovery and mortality reduction was measured. The Catholic University of Honduras Medical Research Group (GIMUNICAH) is working on a more extensive phase 3 trial.

**Trial registration:** ClinicalTrials.gov NCT04771013. February 25, 2021.

## 1. Introduction

Severe Acute Respiratory Syndrome Coronavirus 2 (SARS-CoV-2), responsible for the coronavirus disease 2019 (Covid-19) global pandemic, continues to spread rapidly with increasing daily hospitalizations and deaths worldwide.^1^ Even though vaccine platforms are expected to significantly reduce the burden of disease,^2^ the amount of active Covid-19 cases continue to demand the development of effective treatments. Emerging viral variants ^3,4^ and vaccination hesitancy ^5,6^ may extend or aggravate the problem, particularly in developing countries where access to vaccines is already limited.^7^

The glucocorticoid dexamethasone, alone or in combination with tocilizumab, is the only treatment that has shown a reduction in mortality in large randomized controlled trials, ^8^ a benefit circumscribed to patients requiring oxygen supplementation.^9^ Results from corticosteroids suggest that immune regulation is key in Covid-19 management. Nevertheless, immunosuppression alone might be improved by more complex regulatory mechanisms.

One such theorized approach by the Catholic University of Honduras Medical Research Group (GIMUNICAH), and previously discussed by others, ^10–16^ is through thymic regulation. Evidence supporting this hypothesis is gradually emerging, and the virtual absence of side effects ^10^ confers thymic peptides a particularly advantageous potential in the management of vulnerable groups most affected by Covid-19.

A mechanistic model by Palmer et al. showed that the risk of Covid-19 hospitalization rises exponentially with age and inversely proportional to T-cell production and thymus degeneration. ^17^ In an observational study conducted on ICU patients, enlargement of the thymus during infection was associated with lower mortality rates. ^18^ Further research on *ex vivo* treatment with thymosin alpha 1 (Tα1) of blood from Covid-19 patients has revealed mitigation of cytokine expression and inhibited lymphocyte activation in a CD8+ T-cell subset. ^19^ Finally, retrospective studies of subcutaneous Tα1 and a prospective randomized controlled trial on intramuscular thymic peptides in Covid-19 patients have shown reduction in mortality rates and improvement of the T cell system with normalization of IL-6 levels, respectively. ^20–22^

The pathogenesis of Covid-19 is thought to be driven by SARS-COV-2 replication in the early stages of disease, while a dysregulated immune/inflammatory response appears to promote tissue damage in later phases.^8^ Therefore, the use of thymic peptides might improve immunomodulation and clinical outcomes in the complex management of COVID-19 of moderate to severe cases. We report a nonrandomized phase 2 clinical trial with historic controls from Hospital Santa Bárbara Integrado (HSBI) to evaluate the safety and efficacy of oral thymic peptides in the treatment of hospitalized Covid-19 patients in Honduras.

## 2. Methods

### 2.1 Trial design and oversight

We conducted a nonrandomized, open-label, phase 2 clinical study to evaluate the safety and efficacy of thymic peptides in the treatment of hospitalized Covid-19 patients in Honduras. All patients were enrolled from HSBI in consecutive sampling. A participant-level comparison based on a control group by propensity score matching from registry data was performed.

Hospitalized patients who were 21 years of age or older (with no upper age limit) with confirmed Covid-19 by detection of viral nucleic acid (RNA), viral antigen, or antibodies to the virus were eligible for enrollment. All participants were Stage IIb under the Honduran Ministry of Health Guidelines for Clinical Management of Covid-19 for Adult Patients, ^23^ defined as a patient with or without risk factors that presents with warning signs (shortness of breath, tachypnea), and altered inflammatory parameters. Patients were required to present with at least one of the following: oxygen saturation level below 94%; complete blood count showing lymphopenia, neutrophilia, or both; positive C-reactive protein; chest radiography or computed tomography scan with ground-glass opacities. All patients were hospitalized with oxygen by mask or nasal prongs, which corresponds to a score of 5 according to the World Health Organization (WHO) clinical progression scale. ^24^ Pregnant and breast-feeding women, as well as organ transplant recipients, were not eligible. For the comparison group, registry data from June 2020 to February 2021 was considered, as standardization of Stage IIB therapeutic management to its current guideline in Honduras occurred on May 2020. ^23^

### 2.2 Procedures

Thymic peptides were isolated from 25 thymus glands of 6- to 10-month-old calves bred in an organic production system (free of antibiotics, antiparasitic drugs, anabolic steroids, or other hormonal treatment), through acid lysis with a modified protocol from GIMUNICAH, prepared by the Universidad Católica de Honduras. The entire cervical and thoracic thymus glands portions were utilized, obtaining 100g of lyophilized product. Crystalized stevia was used as sweetener. The amino acidic composition of the calf thymus lysate, as determined by reversed-phase high-performance liquid chromatography after hydrolysis, is detailed in Table S1. Thymic peptides were administered orally once a day by trained physicians either one hour before or two hours after a meal, in a 250-mg dose, dissolved in 50 mL of water, until hospital discharge or death within a 20-day period.

Patients were evaluated daily during their hospitalization, from day 1 through 20. Clinical status was assessed on an eight-category ordinal scale (defined below). Adverse events ≥ Grade 3, as defined by the Common Terminology Criteria for Adverse Events Version 5.0 (CTCAE v5.0), were analyzed. The occurrence of side effects was assessed weekly through the General Assessment of Side Effects (GASE) form. Patients who agreed to follow-ups by phone communication were monitored for adverse events or side effects up to two weeks after discharge.

### 2.3 Outcomes

The primary outcome measures included time to recovery and the number of participants with treatment-related adverse events and side effects. The first was measured in days to clinical recovery, defined as the first day, during the 20 days after enrollment, on which a patient met the criteria for category 1, 2, or 3 on an eight-category ordinal scale (as described by Beigel *et al*.). ^25^ The categories are as follows: 1, not hospitalized and no limitations of activities; 2, not hospitalized, with limitation of activities, home oxygen requirement, or both; 3, hospitalized, not requiring supplemental oxygen and no longer requiring ongoing medical care; 4, hospitalized, not requiring supplemental oxygen but requiring ongoing medical care (related to Covid-19 or other medical conditions); 5, hospitalized, requiring any supplemental oxygen; 6, hospitalized, requiring noninvasive ventilation or use of high-flow oxygen devices; 7, hospitalized, receiving invasive mechanical ventilation or extracorporeal membrane oxygenation; and 8, death. The time to recovery assessment was modified from the first protocol version to better adhere to WHO recommendations on clinical progression outcomes. An amendment submission was presented to the IRB on April 4, 2021, and it was approved April 8, 2021. Adverse events ≥ Grade 3 were registered using the CTCAE v5.0 and side effects were evaluated as defined by the GASE.

The secondary outcome measure was overall survival, defined as the time from the start of treatment until death due to any reason in the 20-day period. Although hospital average length of stay was removed as a secondary outcome for considering it a less reliable metric, it was analyzed as a complementary analysis using the Kaplan-Meier method.

### 2.4 Statistics

For the comparison group, propensity score matching using IBM SPSS ver.25 (IBM Co., Armonk, NY, USA) was performed based on registry data from HSBI. Heatmap and dimensional reduction techniques using principal component analysis were applied to determine the two groups global comparison.

Analyses on time to recovery, mortality, length of stay, and time to supplemental oxygen withdrawal were estimated using the Kaplan–Meier method. Cumulative incidence curves were compared between the two groups with the log-rank test. The Cox proportional-hazard model was used to estimate the hazard ratio and 95% confidence interval. For time to recovery, data for patients who died or did not recover were censored at day 20. For mortality, patients who did not die were censored at day 20. For length of stay, patients who were not discharged at day 20 and patients who died were censored at day 20. For supplemental oxygen withdrawal, patients who still required oxygen therapy after day 20 and patients who died were censored at day 20.

Differences in base drug treatments among groups were analyzed using the Chi-square test or the Fisher’s exact test when appropriate. Safety analysis findings are descriptive in nature and not based on formal statistical hypothesis testing. The number of patients that presented adverse events in the prospectively treated group which were ≥ Grade 3 according to CTCAE v5.0, and the number that manifested side effects according to GASE, were considered. All P values are two-sided, and all analyses were performed according to the intention-to-treat principle.

### 2.5 Ethics approval

Written informed consent prior to inclusion in the study was obtained from all participants or a legal representative if they were unable to provide it. Research was conducted according to the Declaration of Helsinki principles. The trial protocol was approved by the Catholic University of Honduras IRB in Tegucigalpa. The study was approved and registered by the national regulatory entity for clinical trials, the General Directorate for Regulatory Framework Surveillance of the Ministry of Health of Honduras (DGVMN) the eighth of February of 2021; enrollment began tenth of February of 2021. The IRB did not approve prospective controls for this phase 2 trial but authorized a historic control group. Treatment approval for the clinical trial was given until 20 patients in the group receiving oral thymic peptides reached conclusion either by medical discharge or death within a 20-day hospitalization follow-up. Data collection approval was granted for safety data follow-up of discharged patients, which occurred by phone communication for up to two more weeks.

### 2.6 Role of the Funding source

The Universidad Católica de Honduras had no role in the design or conduct of the study; collection, management, analysis, and interpretation of the data; preparation, review, or approval of the manuscript; and decision to submit the manuscript for publication.

## 3. Results

### 3.1 Patients

Between February 10, 2021, and April 12, 2021, a total of 31 patients were prospectively assessed for eligibility for the intervention group (Fig. 1). Of these patients, 22 consented to participate (for the intention-to-treat population) and 20 completed the 20-day trial intervention. Two patients decided to self-discharge against medical advice after manifesting clinical improvement. Out of the 20 remaining patients, 18 recovered during the follow-up period.

**Fig. 1.**
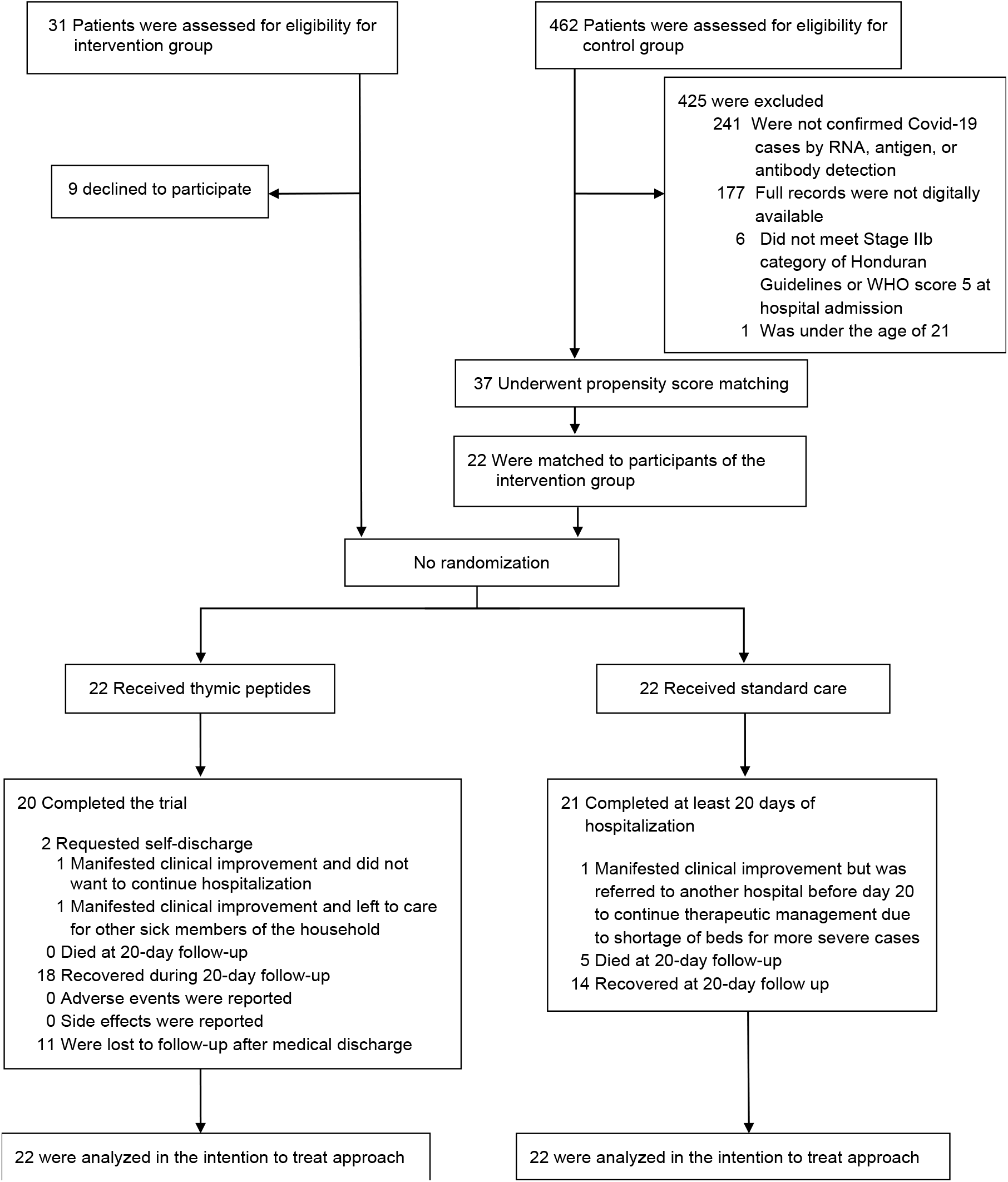
Enrollment and propensity score matching.

For the comparison group, 462 patient records from the Covid-19 hospital ward at HSBI were assessed for eligibility (Fig. 1). Records were filtered in the hospital database by study inclusion and exclusion criteria and digital completeness. Researchers were blind to patient outcomes at this point. Covid-19 confirmatory tests were missing for 241 patient records, and 177 were not digitally available. Stage IIb criteria of Honduran guidelines or WHO score 5 of clinical progression scale were not met for six patients, and one was under the age of 21. The 37 full files of the remaining patients were requested to the statistics department of the hospital for propensity score matching (PSM).

The 22 patients that received thymic peptides and the 37 patients from registry data who received standard care underwent PSM for known Covid-19 severity variables (Table S2). ^26^ The lowest match tolerance (MT = 0.4) was used in SPSS ver.25 that allowed for 22 paired patients. The characteristics of the resulting sample of 44 patients for the intention-to-treat analysis are shown in Table 1. There was a significant (P=0.01) difference in the percentage of diabetic patients between the thymic peptides group (50%) and the standard care group (13.6%), but there were no significant differences between the groups in any other baseline characteristic. Dimensionality reduction by principal component analysis (PCA) demonstrates the overlapping of both groups, and heatmap analysis shows a homogeneous behavior on baseline characteristics (Fig. 2). Together, these results indicate that groups were globally similar on severity indices after matching.

**Table 1.** Demographic and clinical characteristics of the patients at baseline. ^a^.

| Characteristic | Thymic peptides (N=22) | Standard care (N=22) | Total (N=44) |
| --- | --- | --- | --- |
| Age |  |  |  |
| Mean -yr | 52±16 | 57±17 | 54±16 |
| Distribution - no. (%) |  |  |  |
| ≤60 | 15 (68.2) | 12 (54.5) | 27 (61.4) |
| 61-64 | 2 (9.1) | 3 (13.6) | 5 (11.4) |
| ≥ 65 | 5 (22.7) | 7 (31.8) | 12 (27.3) |
| Sex – no. (%) |  |  |  |
| Female | 7 (31.8) | 9 (40.9) | 16 (36.4) |
| Male | 15 (68.2) | 13 (59.1) | 28 (63.6) |
| Race - no. (%) <sup>b</sup> |  |  |  |
| Mestizo | 22 (100) | 22 (100) | 44 (100) |
| Median no. of days since symptom onset (IQR) | 11.5 (9-13) | 10.0 (9-14) | 10.5(9-13) |
| No. of comorbidities | 2±1 | 2±2 | 2±1 |
| Coexisting conditions - no. (%) |  |  |  |
| Diabetes <sup>c</sup> | 11 (50) | 3 (13.6) | 14 (31.8) |
| Hypertension | 7 (31.8) | 9 (40.9) | 16 (36.4) |
| Obesity | 2 (9.1) | 4 (18.2) | 6 (13.6) |
| COPD | 1 (4.5) | 1 (4.5) | 2 (4.5) |
| Heart Failure | 0 (0) | 1 (4.5) | 1 (2.3) |
| Organ damage (other than lung) <sup>d</sup> | 12 (54.5) | 15 (68.2) | 27 (61.4) |
| WHO Clinical Progression Score 5 - no. (%) | 22 (100) | 22 (100) | 44 (100) |
| Heart rate distribution- no. (%) |  |  |  |
| 51-90 | 14 (63.6) | 10 (45.5) | 24 (54.5) |
| 91-110 | 6 (27.3) | 10 (45.5) | 16 (36.4) |
| 111-130 | 2 (9.1) | 2 (9.1) | 4 (9.1) |
| Systolic blood pressure distribution- no. (%) |  |  |  |
| 90-219 | 22 (100) | 22 (100) | 44 (100) |
| Respiratory rate distribution- no. (%) |  |  |  |
| 21-24 | 2 (9.1) | 3 (13.6) | 5 (11.4) |
| ≥ 25 | 20 (90.9) | 19 (86.4) | 39 (88.6) |
| Oxygen saturation distribution- no. (%) |  |  |  |
| 92-93 | 3 (13.6) | 3 (13.6) | 6 (13.6) |
| ≤91 | 19 (86.4) | 19 (86.4) | 38 (86.4) |
| Temperature distribution- no. (%) |  |  |  |
| 35.6-37.9 | 19 (86.4) | 19 (86.4) | 38 (86.4) |
| 38-39 | 3 (13.6) | 2 (9.1) | 5 (11.4) |
| ≥39.1 | 0 (0) | 1 (4.5) | 1 (2.3) |
| SARS-CoV-2 positive test result- no. (%) | 22 (100) | 22 (100) | 44 (100) |

a. Plus-minus values are means ±SD. Percentages may not total 100 because of rounding. IQR denotes interquartile range, COPD Chronic obstructive pulmonary disease, WHO World Health Organization, and SARS-CoV-2 severe acute respiratory syndrome coronavirus 2.
b. Race was recorded in the patient's electronic health record.
c. There was a significant (P=0.01) difference in the percentage of diabetic patients between the thymic peptides group and the standard care group, but there were no significant differences between the groups in any other baseline characteristic.
d. Liver damage was defined as elevated liver enzymes, and kidney damage as elevated creatinine level.

**Fig. 2.**
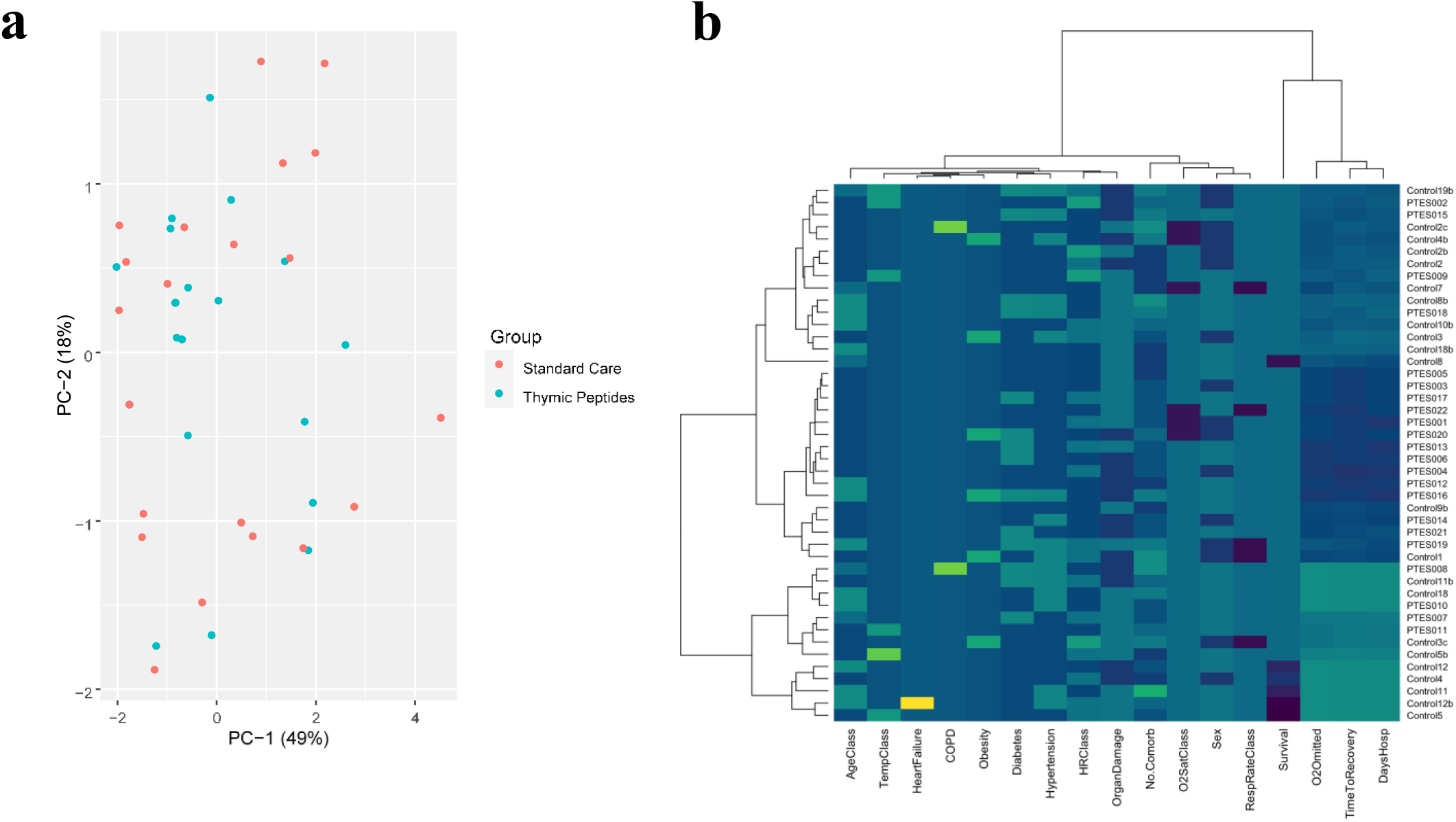
Principal component analysis (PCA) and hierarchical heat map cluster of group characteristics. The percentage of variation explained by each component is indicated in parenthesis in panel A. PCA considered the following variables: confirmatory covid test, number of comorbidities, sex, WHO clinical progression score, need for oxygen therapy, age distribution, heart rate distribution, systolic blood pressure distribution, respiratory rate distribution, oxygen saturation distribution, temperature distribution, and presence of diabetes, hypertension, obesity, overweight, chronic obstructive pulmonary disease, heart failure, organ damage, and dyspnea. Hierarchical heat map cluster of baseline patient characteristics and outcomes is shown in panel B. AgeClass indicates age distribution, TempClass temperature distribution, COPD Chronic obstructive pulmonary disease, HRClass heart rate distribution, No.Comorb number of comorbidities, O2SatClass oxygen saturation distribution, RespRateClass respiratory rate distribution, O2Omitted time to oxygen withdrawal by day 20, DaysHosp time to discharge by day 20.

The median number of days between symptom onset and hospitalization/enrollment was 11.5 (interquartile range, 9 to 13) in the intervention group, and 10 (interquartile range, 9 to 14) in the standard care group. In the thymic peptides and standard care groups, 15 (68.2%) and 13 (59.1%) were men, respectively, and the mean (±SD) age was 52±16 years and 57±17 years, respectively. All patients were mestizo. Most patients had one or more comorbidities (75%), mainly hypertension (36.4%) and diabetes (31.8%). Of note, 61.4% of patients in both groups had elevated liver enzymes or creatinine levels at admission. All patients were WHO clinical progression score 5 at hospitalization/enrollment, and most (86.4%) had an oxygen saturation of ≤91%.

### 3.2 Clinical Management Before and During Hospitalization

Although highly controversial among the Honduran scientific community, guidelines approved by the Honduran Ministry of Health for the treatment of Covid-19 include the use of ivermectin, azithromycin, zinc, hydroxychloroquine or chloroquine, acetaminophen, ibuprofen, and oral disinfectants (sodium hypochlorite, hypochlorous acid, hydrogen peroxide, among others) for the initial phases of disease progression. ^23^ On top of these, therapeutic options for patients presenting with pulmonary compromise before or during hospitalization for phase IIb include colchicine, dexamethasone or methylprednisolone, and anticoagulation (rivaroxaban, apixaban, heparin, or enoxaparin). ^23^

Data for therapies used before hospitalization/enrollment was missing for 14 (31.8%) of patients. Of the 30 (68.2%) patients with available information, 14 (46.7%) had used ivermectin, 19 (63.3%) dexamethasone, 13 (43.3%) colchicine, 15 (50%) azithromycin, 15 (50%) hypochlorous acid rinses, 14 (46.7%) zinc, 2 (6.7%) enoxaparin, and 7 (23.3%) rivaroxaban, with no statistically significant difference between groups.

During hospitalization, all patients received dexamethasone and enoxaparin. No patient was administered ivermectin or used hypochlorous acid rinses in the thymic peptides group. In the standard care group, 9 (40.9%) patients used ivermectin, and 14 (63.6%) used hypochlorous acid rinses. Significant differences were observed between the intervention and control groups, showing a reduced use of colchicine (45.5% vs 95.5%, P<0.001), azithromycin (9.1% vs 68.2%, P<0.001), zinc (9.5% vs 72.7%, P<0.001), and ceftriaxone (40.9% vs 86.4%, P = 0.02) by the thymic peptides group. Nevertheless, significantly more patients received vitamin D during hospitalization in the thymic peptides group compared to standard of care (95.5% vs. 40.9%, P<0.001). It is important to note that one patient received hydroxychloroquine in the intervention group, but as continuation of her rheumatoid arthritis chronic management, not as Covid-19 treatment.

### 3.3 Primary outcomes

#### 3.3.1 Time to Recovery

Heatmap clusters showed differences between group behavior in time to recovery (Fig. 2). The median time to recovery in the thymic peptides group was 6 days, as compared with 12 days of standard care (Table S3). The Kaplan-Meier analysis revealed a significantly shorter time to recovery in the intervention group (log-rank test of P=0.002) (Fig. 3). The hazard ratio for recovery was 2.75, with a 95% confidence interval (CI) of 1.34 to 5.62.

**Fig. 3.**
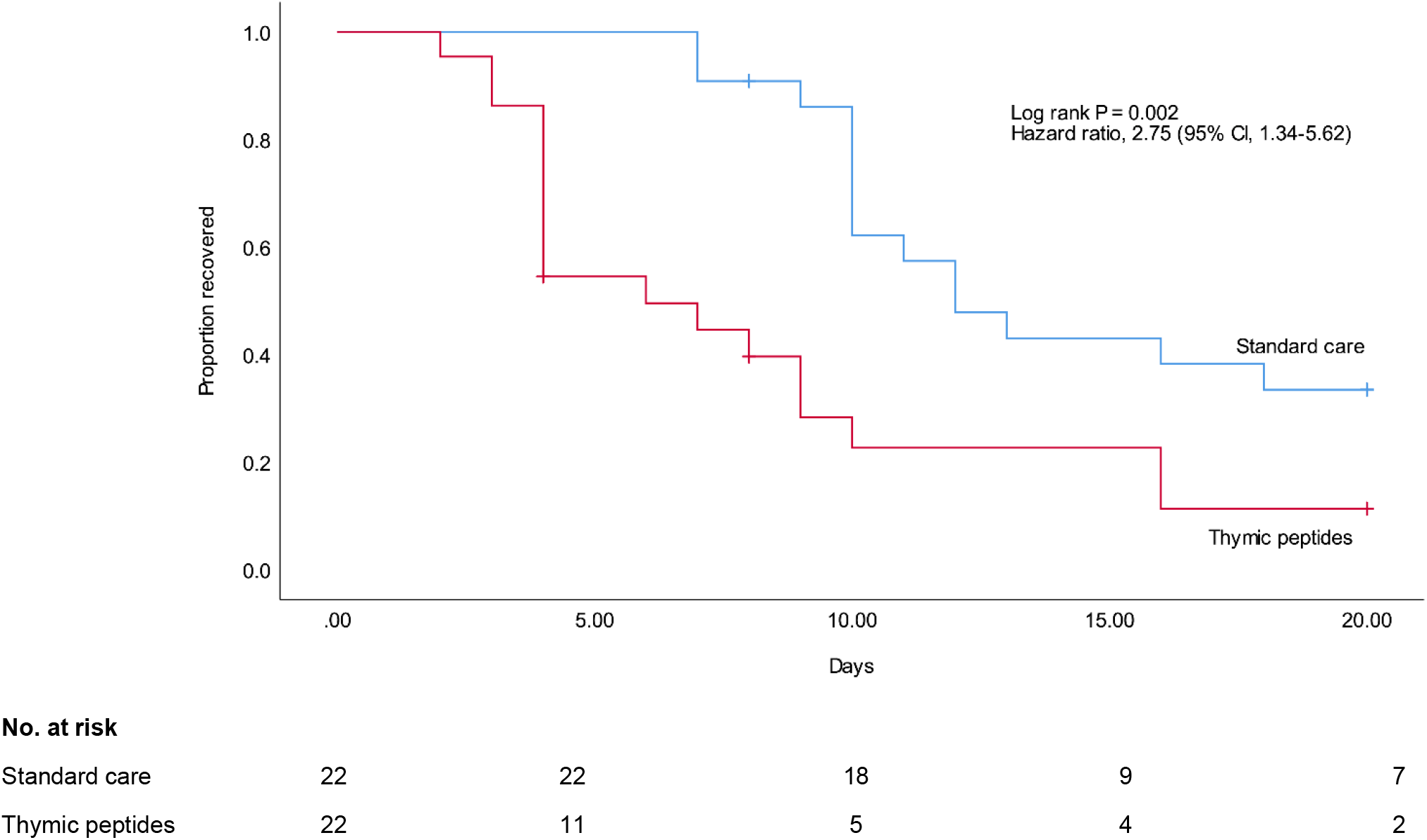
Kaplan-Meier estimates of time to recovery by day 20. The Kaplan-Meier method was used to estimate the cumulative proportion of patients and the log-rank test was used to compare the two groups. The Cox proportional-hazard model was used to estimate the hazard ratio and 95% confidence interval. Vertical dashes indicate censored data.

### 3.3.2 Safety

The GASE instrument was evaluated weekly by physicians of the Covid-19 ward for every patient in the thymic peptides group. No side effect related to thymic peptides was reported during the 20-day follow-up hospitalization period. Adverse events ≥ Grade 3 as defined by the CTCAE v5.0 were monitored daily for every patient. No adverse event was reported during the 20-day follow-up hospitalization period. After the 20-day intervention cutoff, side effects or adverse events were monitored for up to two more weeks. Discharged patients were interviewed to the cellphone number reported in the patient’s file. However, 11 patients were lost to follow-up after medical discharge. For patients monitored, no side effects or adverse events related to thymic peptides were reported.

### 3.4 Secondary outcomes

#### 3.4.1 Mortality

Heatmap clusters also showed differences between group behavior in mortality (Fig. 2). The
re were no deaths in the thymic peptides group by day 20. In contrast, the Kaplan-Meier estimate of mortality for the standard care group by day 20 was 24% (Fig. 4). This difference was statistically significant (log-rank P=0.02). The log-rank statistic was used alone because the Cox proportional-hazard model results in a degenerate estimation for the group with no events.

**Fig. 4.**
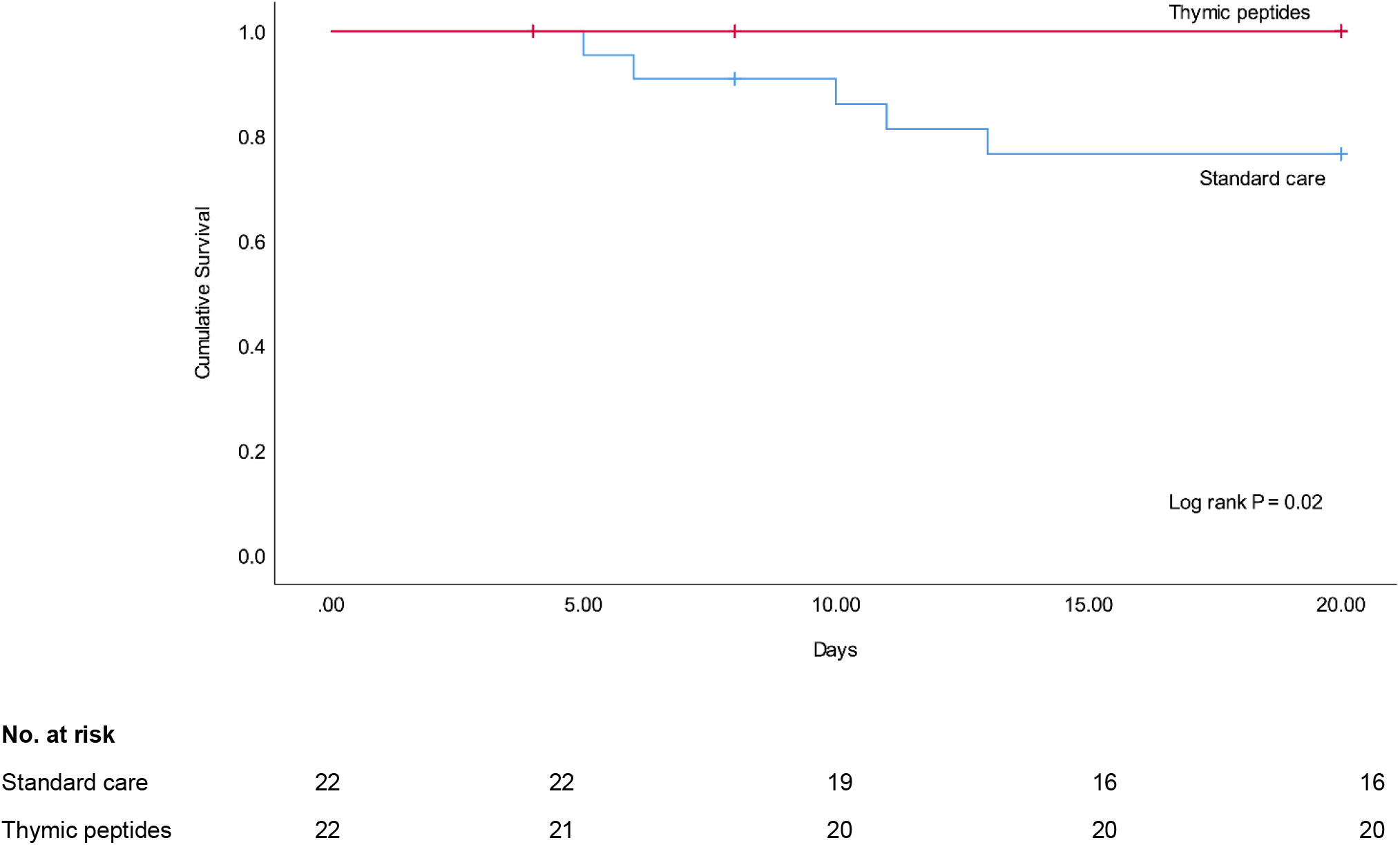
Kaplan-Meier estimates of mortality by day 20. The Kaplan-Meier method was used to estimate the cumulative proportion of patients and the log-rank test was used to compare the two groups. Vertical dashes indicate censored data

#### 3.4.2 Other Clinical Outcomes

Kaplan-Meier analysis showed a median time to oxygen therapy withdrawal of 4 days in the thymic peptides group, as compared with 10 days in standard care (hazard ratio, 2.3; 95% CI, 1.13 to 4.67; log-rank P=0.01) (Fig. S1 and Table S4). Finally, patients in the intervention group had a shorter length of in-hospital stay (median, 6 days, as compared with 12 days; hazard ratio for discharge, 2.34; 95% CI, 1.16-4.75; log-rank P=0.01) (Fig. S2 and Table S5). These differences in the previous outcomes can be visualized in the heatmap hierarchical clusters (Fig. 2).

## 4. Discussion

This nonrandomized phase 2 clinical trial of thymic peptides for the treatment of hospitalized Covid-19 patients reported no side effects, adverse events, or deaths by the 20-day cutoff point. In addition, when compared with registry patient data after PSM, a significantly shorter time to recovery (median, 6 days vs. 12 days; hazard ratio for recovery, 2.75 [95% CI, 1.34 to 5.62]) and mortality reduction (log-rank P=0.02) was observed. This is the first clinical trial of oral thymic peptides in the management of Covid-19 patients.

Only one other prospective study has evaluated an administration form of thymic peptides in hospitalized Covid-19 patients. Khavinson et al. analyzed the addition of intramuscular thymic peptides to standard therapy. They found an accelerated reduction of pro-inflammatory markers, as well as improvement in T cell system indicators. ^22^ No deaths were reported in any of the groups. ^22^ However, safety data and time to recovery were not described.

Retrospective evidence on thymic peptides in Covid-19 therapy can be found for subcutaneous Tα1 alone. Consistent with our trial, a study by Wu et al. found a significant decrease in 28-day mortality of the Tα1 group relative to the non-thymosin control (7.8% vs. 14.7%, P = 0.016). ^20^ Prolonged hospital length of stay and disease duration were described; although this corresponded to critical type patients. ^20^ No safety data was included. Liu et al. also reported a significant reduction in mortality of severe Covid-19 patients who used this specific peptide (11.11% vs. 30%, P=0.04). ^21^ They found no side effects related to the treatment. Finally, Sun et al. showed a significantly lower 28-day mortality in the Tα1 group relative to control (41.3% vs. 60.6%, P < 0.001), but after adjusting by baseline confounders, the difference became non-significant (51.0% vs. 52.9%). ^27^ No safety data was described in this study either.

The results with Tα1 are suggestive of the benefits that this immunomodulatory approach can add to Covid-19 therapy. However, peptides combination rather than isolated pharmacological agents might enhance the clinical response through synergism. A recent preclinical study of recombinant human thymosin beta-4 in mice found a significantly increased survival rate by inhibiting viral replication, modulating the immune response, and promoting tissue repair. ^28^ Thymulin is another nonapeptide secreted by thymic epithelial cells with essential regulatory activities. It has been shown to suppress p38, a MAPK family member implicated in glucocorticoid resistance, and inhibit NF-kB activation, with crucial pathogenic effects in lung disease. ^29^

Limitations of our study include its small sample size and the lack of a prospectively randomized control group. However, valuable information was acquired through PSM of registry data that accounts for standard care results in a group of patients globally similar in baseline characteristics of possible severity confounders. Also, poor socioeconomic conditions could have influenced the low remote cellphone follow-up response rate after discharge (55%) by lack of patient access to internet or a service plan. A phase 3 trial by the GIMUNICAH group is underway to address the need for confirmatory analyses in a larger population. This was a single-center study that included only mestizo population, which might have impacted its universal validity. Nonetheless, given the underrepresentation of this population in Covid-19 clinical trials, data obtention is highly relevant. ^30^

In conclusion, given that a dysregulated immune/inflammatory response promotes tissue damage in later phases of COVID-19,^8^ the use of thymic peptides should be further examined and considered to improve immunomodulation and clinical outcomes in the complex management of the disease. The daily oral dose of 250 mg of thymic peptides proved to be safe in a hospitalized Covid-19 group of patients in Honduras, reporting no deaths by day 20. When compared with registry data after PSM, shorter times to oxygen therapy withdrawal, recovery, and length of stay, as well as reduction in mortality were identified. Trials with larger populations are required to confirm these findings. The GIMUNICAH group is already working on a more extensive Phase 3 clinical trial of oral thymic peptides in Covid-19 patients.

## Data Availability

Plan to Share Individual Participant Data (IPD):   Yes
Plan Description:  All of the individual participant data collected during the trial, after deidentification.
Supporting Materials:  Study Protocol
Supporting Materials:  Statistical Analysis Plan (SAP)
Supporting Materials:  Informed Consent Form (ICF)
Supporting Materials:  Clinical Study Report (CSR)
Supporting Materials:  Analytic Code
Time Frame:   Beginning 3 months and ending 5 years following article publication.
Email addresses
Access Criteria:   Investigators whose proposed use of the data has been approved by an independent review committee ("learned intermediary") identified for this purpose.
For individual participant data meta-analysis.

## Acknowledgements

We thank all the patients and their families who participated in this trial. We also acknowledge the contributions of the following nursing staff at Hospital Santa Bárbara Integrado: Edil Eduardo Castellanos Rodríguez, Cristian Omar Pereira, Jessica Membreño, David Armando Cardona, Lesly Xiomara Rodríguez, Yanidia Navidad Reyes Toro, Cinthia Karina Romero, Sindy Verónica Vega Toro, Maryoli Judith Rivera, Susseiry Portillo, Carmen Julissa Leiva, Iris Maribel Lemuz, Rosy Gissela Mejía, Lucy Margoth Pineda, Elsy Maritza Castellanos, Elsy Yolany Arriaga, Yissel Alejandra Barahona, Cesia María Ramos, Alex Alberto Fino, Vicky Soliman, and Nadia Villanueva. We acknowledge ANID BECAS/DOCTORADO NACIONAL 21211334. We also thank Jeimy Paola Reyes Caballero for assistance with patient records processing for PSM; Orfy Yamileth Arita Henriquez for search on thymic peptides formulations; German Reyes for coordination of software communication; Fredy Antonio Guillén Guevara for coordination with the Ministry of Health in Honduras.

## Author contributions

### Authors who have accessed verified the underlying data

Héctor M. Ramos-Zaldívar, Karla Giselle Reyes Perdomo, Nelson A. Espinoza Moreno.

### Study concept and design

Ramos-Zaldívar, Reyes-Perdomo, Espinoza-Moreno, Dox-Cruz, Aguirre Urbina, Rivera Caballero, Perdomo Dominguez, Peña Calix, Monterroso-Reyes, Caballero Vásquez, Zelaya Ortiz, Rodríguez Machado, Forgas Solis, Sebilla Silva, Zavala Galeano, Morga Alvarado, Solís Medina, Guerrero-Díaz, Jiménez-Faraj, Perelló Santos, Moncada Arita, Valdiviezo Montufar, Hernández Sabillón, Sorto G., Padilla Navarro, Palomo Bermúdez, Alvarenga Andino.

### Acquisition, analysis, or interpretation of data

All authors.

### Writing Committee

Ramos-Zaldívar, Reyes-Perdomo, Espinoza-Moreno, Dox-Cruz, Aguirre Urbina, Caballero Vásquez, Peña Calix, Zelaya Ortiz, Rodríguez Machado, Monterroso-Reyes, Rivera Caballero, Sebilla Silva, Morga Alvarado, Padilla Navarro, Solís Medina, Forgas Solis, Moncada Arita, Perdomo Dominguez, Zavala Galeano, Jiménez-Faraj, Palomo Bermúdez, Perelló Santos, Alvarenga Andino, Guerrero-Díaz, Montes-Gambarelli.

### Critical revision of the manuscript for important intellectual content

All authors.

### Statistical analysis

Ramos-Zaldívar, Reyes-Perdomo, Espinoza-Moreno, Andia, Salas-Huenuleo.

### Study supervision

Ramos-Zaldívar, Reyes-Perdomo, Espinoza-Moreno, Reyes Guzman, Rivera Reyes.

All authors had full access to the full data in the study and accept responsibility to submit for publication.

### Funding/Support

The research reported was funded by the Universidad Católica de Honduras.

### Declaration of interests

Héctor M. Ramos Zaldívar, Karla G. Reyes-Perdomo, and Héctor Armando Alvarenga Andino designed the modified protocol for isolation of thymic peptides, which is patent pending. Héctor M. Ramos Zaldívar receives a grant for education from the Universidad Católica de Honduras. The rest of the authors declare no competing interests.

## Availability of data and material

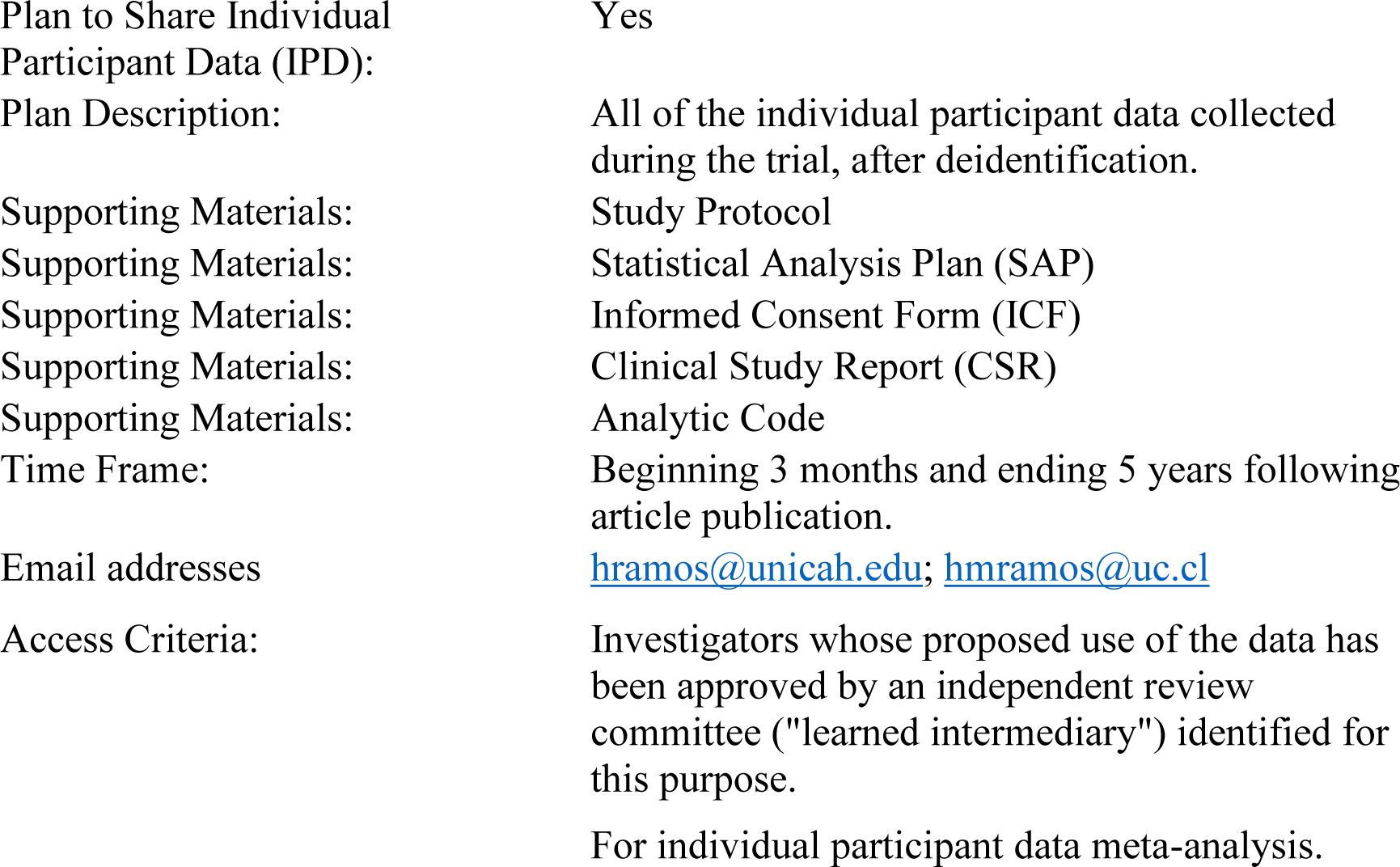

## Notes

### Clinical Trial

ClinicalTrials.gov Identifier: NCT04771013

### Author Declarations

The Catholic University of Honduras IRB in Tegucigalpa gave approval for this work. The study was approved and registered by the national regulatory entity for clinical trials, the General Directorate for Regulatory Framework Surveillance of the Ministry of Health of Honduras (DGVMN) the eighth of February of 2021; enrollment began tenth of February of 2021.

## References

1 COVID-19 Map - Johns Hopkins Coronavirus Resource Center. https://coronavirus.jhu.edu/map.html (accessed May 30, 2021).

2 Tenforde MW, Olson SM, Self WH, et al. Effectiveness of Pfizer-BioNTech and Moderna Vaccines Against COVID-19 Among Hospitalized Adults Aged ≥65 Years - United States, January-March 2021. MMWR Morb Mortal Wkly Rep 2021; 70: 674–9.

3 Hacisuleyman E, Hale C, Saito Y, et al. Vaccine Breakthrough Infections with SARS-CoV-2 Variants. N Engl J Med 2021; published online April. DOI:10.1056/NEJMoa2105000.

4 Wang Z, Schmidt F, Weisblum Y, et al. mRNA vaccine-elicited antibodies to SARS-CoV-2 and circulating variants. Nature 2021; 592: 616–22.

5 Biswas N, Mustapha T, Khubchandani J, Price JH. The Nature and Extent of COVID-19 Vaccination Hesitancy in Healthcare Workers. J Community Health 2021; : 1–8.

6 Daly M, Robinson E. Willingness to Vaccinate Against COVID-19 in the U.S.: Representative Longitudinal Evidence From April to October 2020. Am J Prev Med 2021; 60: 766–73.

7 Choi EM. COVID-19 vaccines for low-and middle-income countries. Trans R Soc Trop Med Hyg 2021; 115: 447–56.

8 COVID-19 Treatment Guidelines Panel. Coronavirus Disease 2019 (COVID-19) Treatment Guidelines. National Institutes of Health. https://www.covid19treatmentguidelines.nih.gov/therapeutic-management/ (accessed June 1, 2021).

9 Horby P, Lim WS, Emberson JR, et al. Dexamethasone in Hospitalized Patients with Covid-19. N Engl J Med 2021; 384: 693–704.

10 Khavinson V, Linkova N, Dyatlova A, Kuznik B, Umnov R. Peptides: Prospects for Use in the Treatment of COVID-19. Molecules 2020; 25. DOI:10.3390/molecules25194389.

11 Rodriguez Martin RR, Gonzalez Gonzalez O, Rodriguez Gonzalez C, Rodriguez Gonzalez RR. Biomodulina T (InmunyVital(®)) Restores T Cells and Helps Contain COVID-19. Front Immunol 2020; 11: 606447.

12 Kellogg C, Equils O. The role of the thymus in COVID-19 disease severity: implications for antibody treatment and immunization. Hum Vaccin Immunother 2021; 17: 638–43.

13 Wang W, Thomas R, Oh J, Su D-M. Thymic Aging May Be Associated with COVID-19 Pathophysiology in the Elderly. Cells 2021; 10. DOI:10.3390/cells10030628.

14 Scarpa R, Costa L, Del Puente A, Caso F. Role of thymopoiesis and inflammaging in COVID-19 phenotype. Pediatr. Neonatol. 2020; 61: 364–5.

15 Rehman S, Majeed T, Ansari MA, Ali U, Sabit H, Al-Suhaimi EA. Current scenario of COVID-19 in pediatric age group and physiology of immune and thymus response. Saudi J. Biol. Sci. 2020; 27: 2567–73.

16 Güneş H, Dinçer S, Acıpayam C, Yurttutan S, Özkars MY. What chances do children have against COVID-19? Is the answer hidden within the thymus? Eur J Pediatr 2021; 180: 983–6.

17 Palmer S, Cunniffe N, Donnelly R. COVID-19 hospitalization rates rise exponentially with age, inversely proportional to thymic T-cell production. J R Soc Interface 2021; 18: 20200982.

18 Cuvelier P, Roux H, Couëdel-Courteille A, et al. Protective reactive thymus hyperplasia in COVID-19 acute respiratory distress syndrome. Crit Care 2021; 25: 4.

19 Matteucci C, Minutolo A, Balestrieri E, et al. Thymosin Alpha 1 Mitigates Cytokine Storm in Blood Cells From Coronavirus Disease 2019 Patients. Open forum Infect Dis 2021; 8: ofaa588.

20 Wu M, Ji J-J, Zhong L, et al. Thymosin α1 therapy in critically ill patients with COVID-19: A multicenter retrospective cohort study. Int Immunopharmacol 2020; 88: 106873.

21 Liu Y, Pan Y, Hu Z, et al. Thymosin Alpha 1 Reduces the Mortality of Severe Coronavirus Disease 2019 by Restoration of Lymphocytopenia and Reversion of Exhausted T Cells. Clin Infect Dis an Off Publ Infect Dis Soc Am 2020; 71: 2150–7.

22 Khavinson VK, Kuznik BI, Trofimova S V, et al. Results and Prospects of Using Activator of Hematopoietic Stem Cell Differentiation in Complex Therapy for Patients with COVID-19. Stem cell Rev reports 2021; 17: 285–90.

23 Protocolo de Manejo Clínico del Paciente Adulto con COVID-19 según las etapas de la enfermedad en las redes de servicio de Salud - Secretaria de Salud Honduras. http://www.salud.gob.hn/site/index.php/component/edocman/protocolo-de-manejo-clinico-del-paciente-adulto-con-covid-19-segun-las-etapas-de-la-enfermedad-en-las-redes-de-servicio-de-salud (accessed June 3, 2021).

24 A minimal common outcome measure set for COVID-19 clinical research. Lancet Infect Dis 2020; 20: e192–7.

25 Beigel JH, Tomashek KM, Dodd LE, et al. Remdesivir for the Treatment of Covid-19 - Final Report. N Engl J Med 2020; 383: 1813–26.

26 Huespe I, Carboni Bisso I, Di Stefano S, Terrasa S, Gemelli NA, Las Heras M. COVID-19 Severity Index: A predictive score for hospitalized patients. Med. intensiva. 2020; published online Dec. DOI:10.1016/j.medin.2020.12.001.

27 Sun Q, Xie J, Zheng R, et al. The effect of thymosin α1 on mortality of critical COVID-19 patients: A multicenter retrospective study. Int Immunopharmacol 2021; 90: 107143.

28 Yu R, Mao Y, Li K, et al. Recombinant Human Thymosin Beta-4 Protects against Mouse Coronavirus Infection. Mediators Inflamm 2021; 2021: 9979032.

29 Santos M, Henriques-Coelho T, Leite-Moreira A. Immunomodulatory role of thymulin in lung diseases. Expert Opin Ther Targets 2010; 14: 131–41.

30 Chastain DB, Osae SP, Henao-Martínez AF, Franco-Paredes C, Chastain JS, Young HN. Racial Disproportionality in Covid Clinical Trials. N Engl J Med 2020; 383: e59.

