## Supplementary Material for "A nonrandomized phase 2 trial of oral thymic peptides in hospitalized patients with Covid-19"

---

Héctor M. Ramos-Zaldívar, M.D., [1,2] Karla G. Reyes-Perdomo, M.D.,[1,3] Nelson A. Espinoza-Moreno, M.D.,[1] Ernesto Tomás Dox-Cruz,[1] Thania Camila Aguirre Urbina,[1] Astrid Yohaly Rivera Caballero,[1] Eduardo Smelin Perdomo Dominguez,[1] Sofía Guadalupe Peña Calix ,[1] Joselin Michelle Monterroso-Reyes,[1] Erick Fernando Caballero Vásquez,[1] Tarek Sai Zelaya Ortiz,[1] Hilbron Eduardo Rodríguez-Machado,[1] Marcelo Andres Forgas Solis, M.D.,[1] Iveth Sebilla Silva,[1] Mauricio Edgardo Zavala Galeano,[1] Alejandro Antonio Morga Alvarado,[1] Angie María Nicolle Solís Medina,[1] Leticia M. Guerrero-Díaz,[1] Julia E. Jiménez-Faraj, M.D.,[1,4] Caroll Alejandra Perelló Santos, M.D.,[1,5] Wilberg A. Moncada Arita, M.D.,[1] Darwing Fabricio Valdiviezo Montufar,[1] Josué David Hernández Sabillón,[1] Mónica L. Sorto G.,[1] Xochilt Xiomara Padilla Navarro,[1] Victoria A. Palomo-Bermúdez, M.D.,[1,5] Héctor Armando Alvarenga Andino, MSc,[1] Sandra Patricia Reyes Guzman, M.D.,[6] María Haydee Rivera Reyes, M.D.,[6] Esdras Said Medina Paz, M.D.,[6] Joselyn Rosario Alvarado Enamorado, M.D.,[6] Yenny Mariel Sabillón Sagastume, M.D.,[6] Ariadna Stephanny Mejia Rivera, M.D.,[6] Claudia Michelle Posas Sarmiento, M.D.,[6] Xenia Vanessa Jiménez Pineda, M.D.,[6] Verónica Alejandra Hernández Puerto, M.D.,[6] Josué David Portillo Landaverde, M.D.,[6] Sergio Reyes S., M.D.,[6] Ivin Perdomo R., M.D.,[6] Josué J. Rivera, M.D.,[6] Wendy Cecilia Mendoza Girón, M.D.,[6] Karla Melissa Tróchez Sabillón, M.D.,[6] Paola Nohemy Katsumata Leiva, M.D.,[7] Karla Elizabeth Pineda Toro, M.D.,[7] Jimena A. Montes-Gambarelli, M.D.,[1] Cristhiam Flores, P.G.Cert.,[1,8] Edison Salas-Huenuleo, Ph.D.,[9] Marcelo E. Andia, M.D.[10,11]

1. Grupo de Investigación Médica de la Universidad Católica de Honduras, (GIMUNICAH), Faculty of Medicine, Universidad Católica de Honduras.
2. Doctoral Program in Medical Sciences, Faculty of Medicine, Pontificia Universidad Católica de Chile.
3. Postgraduate Program on Breastfeeding and Nutrition in Children Under the Age of Two, Faculty of Health Sciences, Universidad Mayor, Chile.
4. Hospital Civil de Guadalajara Juan I. Menchaca, México.
5. Hospital del Valle, San Pedro Sula, Honduras.
6. Hospital Santa Bárbara Integrado, Santa Bárbara, Honduras.
7. Triaje de Santa Bárbara, Secretaría de Salud de Honduras.
8. Laboratorio de Biología Molecular de San Pedro Sula, Secretaría de Salud de Honduras.
9. Advanced Integrated Technologies, (AINTECH), Santiago, Chile.
10. Biomedical Imaging Center Radiology Department, School of Medicine, Pontificia Universidad Católica de Chile, Santiago, Chile.
11. Millennium Nucleus in Cardiovascular Magnetic Resonance, Santiago, Chile.

Corresponding author: Héctor Miguel Ramos Zaldívar, M.D.

Contact information: Primary

Secondary

ORCID iD: https://orcid.org/0000-0002-0612-8289

### SUPPLEMENTARY INFORMATION

**Supplementary Tables**

**Table S1.**
**Aminoacidic composition of Calf Thymus Lysate as determined by Reversed-Phase High Performance Liquid Chromatography after Hydrolisis.**

| | Amino acid | Micrograms / milliliter | Essential amino acid (E) |
|---|---|---|---|
| 1 | Aspartic acid | 56 | - |
| 2 | Glutamic acid | 73 | - |
| 3 | Alanine | 28 | - |
| 4 | Arginine | 31 | E |
| 5 | Phenylalanine | 28 | E |
| 6 | Glycine | 10 | - |
| 7 | Histidine | 22 | E |
| 8 | Isoleucine | 12 | E |
| 9 | Leucine | 54 | E |
| 10 | Lysine | 66 | E |
| 11 | Methionine | 4 | E |
| 12 | Proline | 15 | - |
| 13 | Serine | 23 | - |
| 14 | Tyrosine | 23 | - |
| 15 | Threonine | 27 | E |
| 16 | Valine | 24 | E |

**Table S2.**
**Variables analyzed for Propensity Score Matching**

| | |
|---|---|
| 1 | Age distribution |
| | ≤60 |
| | 61-64 |
| | ≥ 65 |
| 2 | Sex |
| 3 | Number of comorbidities |
| 4 | Diabetes diagnosis |
| 5 | Hypertension diagnosis |
| 6 | Obesity diagnosis |
| 7 | Overweight diagnosis |
| 8 | Chronic obstructive pulmonary disease diagnosis |
| 9 | Heart failure diagnosis |
| 10 | Organ damage other than lung [a] |
| 11 | WHO clinical progression score |
| 12 | Need for supplemental oxygen |
| 13 | Heart rate distribution |
| | ≤40 |
| | 41-50 |
| | 51-90 |
| | 91-110 |
| | 111-130 |
| | ≥131 |
| 14 | Systolic blood pressure distribution |
| | ≤90 |
| | 90-219 |
| | ≥220 |
| 15 | Respiratory rate distribution |
| | 12-20 |
| | 21-24 |
| | ≥ 25 |
| 16 | Oxygen saturation distribution |
| | ≤91 |
| | 92-93 |
| | 94-95 |
| | ≥96 |
| 17 | Temperature distribution |
| | 35.6-37.9 |
| | 38-39 |
| | ≥39.1 |
| 18 | Presence of dyspnea |

a. Liver damage was defined as elevated liver enzymes, and kidney damage as elevated creatinine level.

**Table S3.**
**Means and Medians for Participant Recovery by Day 20.**

| | Mean[a] | | | | Median | | | |
|---|---|---|---|---|---|---|---|---|
| | | | 95% Confidence Interval | | | | 95% Confidence Interval | |
| **Group** | **Estimate** | **Std. Error** | **Lower Bound** | **Upper Bound** | **Estimate** | **Std. Error** | **Lower Bound** | **Upper Bound** |
| **Standard care** | 14.081 | 1.051 | 12.021 | 16.142 | 12.000 | 1.519 | 9.023 | 14.977 |
| **Thymic peptides** | 8.345 | 1.272 | 5.851 | 10.838 | 6.000 | 1.030 | 3.981 | 8.019 |
| **Overall** | 11.245 | .931 | 9.420 | 13.071 | 10.000 | .510 | 9.000 | 11.000 |

a. Estimation is limited to the largest survival time if it is censored.

**Table S4**.

**Means and Medians for Oxygen Therapy Withdrawal by day 20.**

| Group | Mean[a] | | | | Median | | | |
|---|---|---|---|---|---|---|---|---|
| | Estimate | Std. Error | 95% Confidence Interval | | Estimate | Std. Error | 95% Confidence Interval | |
| | | | Lower Bound | Upper Bound | | | Lower Bound | Upper Bound |
| **Standard care** | 12.692 | 1.284 | 10.175 | 15.209 | 10.000 | 1.476 | 7.106 | 12.894 |
| **Thymic peptides** | 7.730 | 1.302 | 5.177 | 10.283 | 4.000 | 1.642 | .781 | 7.219 |
| **Overall** | 10.220 | .994 | 8.270 | 12.169 | 9.000 | 1.037 | 6.967 | 11.033 |

a. Estimation is limited to the largest survival time if it is censored.

**Table S5.**

**Means and Medians for Length of Hospitalization by day 20.**

| | Mean[a] | | | | Median | | | |
|---|---|---|---|---|---|---|---|---|
| | | | 95% Confidence Interval | | | | 95% Confidence Interval | |
| **Group** | **Estimate** | **Std. Error** | **Lower Bound** | **Upper Bound** | **Estimate** | **Std. Error** | **Lower Bound** | **Upper Bound** |
| **Standard care** | 14.129 | 1.043 | 12.084 | 16.174 | 12.000 | 1.519 | 9.023 | 14.977 |
| **Thymic peptides** | 9.576 | 1.165 | 7.293 | 11.860 | 6.000 | 1.278 | 3.495 | 8.505 |
| **Overall** | 11.904 | .855 | 10.227 | 13.581 | 11.000 | .777 | 9.476 | 12.524 |

a. Estimation is limited to the largest survival time if it is censored.

#### Supplementary Figures

**Figure S1: Kaplan-Meier estimates of time to oxygen therapy withdrawal by day 20.** The Kaplan-Meier method was used to estimate the cumulative proportion of patients and the log-rank test was used to compare the two groups. The Cox proportional-hazard model was used to estimate the hazard ratio and 95% confidence interval. Vertical dashes indicate censored data.

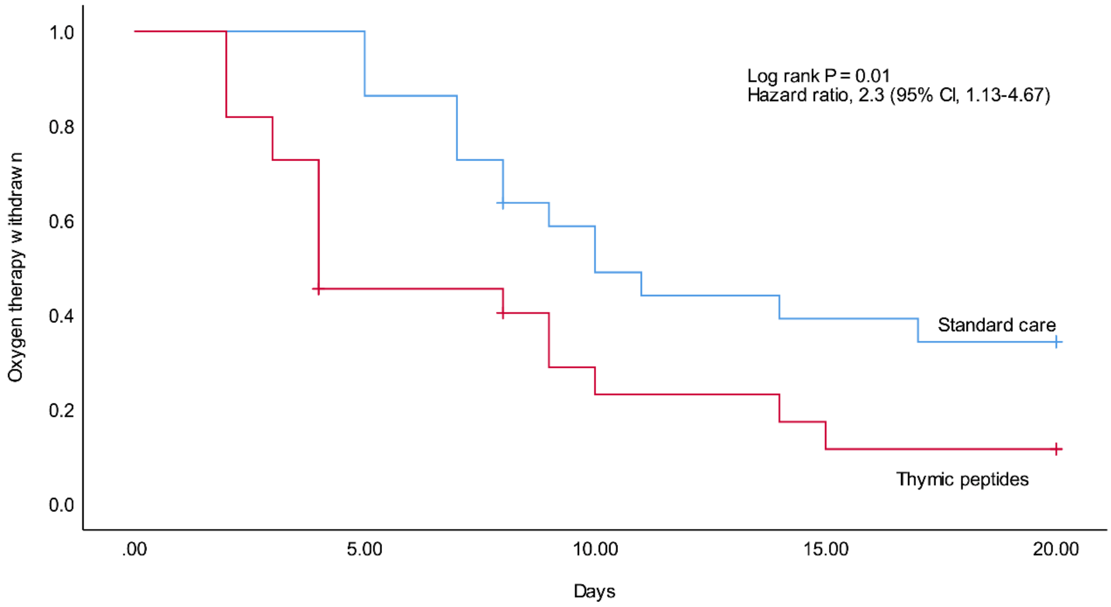

| No. at risk | | | | | |
|---|---|---|---|---|---|
| Standard care | 22 | 22 | 12 | 8 | 7 |
| Thymic peptides | 22 | 9 | 5 | 3 | 2 |

**Figure S2: Kaplan-Meier estimates of time to discharge by day 20.** The Kaplan-Meier method was used to estimate the cumulative proportion of patients and the log-rank test was used to compare the two groups. The Cox proportional-hazard model was used to estimate the hazard ratio and 95% confidence interval. Vertical dashes indicate censored data.

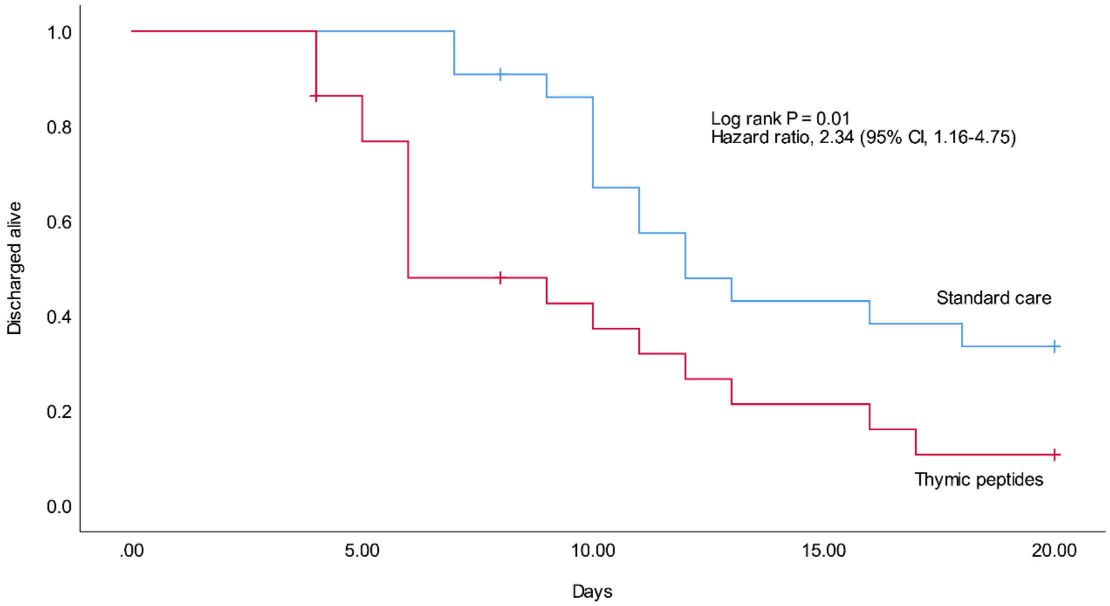

| No. at risk | | | | | |
|---|---|---|---|---|---|
| Standard care | 22 | 22 | 18 | 9 | 7 |
| Thymic peptides | 22 | 18 | 8 | 4 | 2 |
